# The impact of COVID-19 infection on maternal and reproductive health care services in governmental health institutions of Dessie town, North-East Ethiopia, 2020 G.C.

**DOI:** 10.1101/2020.09.20.20198259

**Authors:** Kibir Temesgen, Amare Workie, Tenagnework Dilnessa

## Abstract

**Background:** The COVID 19 pandemic is causing huge stress on the health care system of all countries in the world. The impact of the pandemic is both social and economic. Pregnancy is an exciting and sometimes stressful experience. Being pregnant during a disease outbreak may add extra anxiety and concern for pregnant women and for those who provide care for them [1, 2].

During the initial stages of the pandemic, it appeared Africa would be spared the burden of COVID-19. However, by April 7th, a total of 45 countries within the WHO African region had reported over 7000 cases (although some place it at over 10 000), with at least 292 deaths and 612 people recovered. Ethiopia, being one of the developing countries trying to address the diverse needs of its people, is currently at the verge of the epidemic [5, 7].

**Objectives:** The general objective of this study was to assess the impact of COVID-19 infection on maternal and reproductive health care services among mothers getting service in governmental health institutions of Dessie town, 2020 G.C.

**Methods:** Institution based cross sectional study design using mixed (quantitative supplemented with qualitative) method was employed to identify the impact of COVID-19 infection on maternal and reproductive health care services among women who get service in governmental health institutions of Dessie town. The study was conducted from July 1-15 / 2020.

**Result:** According to this study, Six percent (6%) of antenatal care attendees, 18% of delivery care attendees and nearly half (46.7%) of postnatal care attendees reported inappropriate service delivery due to fear of health care providers, shortage medical supplies and staff work load. The study also showed that utilization of these services was decreased due to fear of clients to go to health institutions.

**Conclusion and recommendation:** This study concluded that COVID-19 significantly affects the quality and utilization of maternal and reproductive health care services. The study also showed that utilization of these services was decreased due to fear of clients to go to health institutions. Ministry of health should continue maternity and reproductive health care services such as family planning to be prioritized as an essential core health service.

## 1. INTRODUCTION

The COVID-19 pandemic has in a matter of weeks fundamentally crippled health systems in many countries and is now threatening to cause a global economic depression. This in turn is having an unprecedented impact on health care organizations’ ability to provide emergency care and on societies to provide core functions of the state. The COVID 19 pandemic is causing huge stress on the health care system of all countries in the world. The impact of the pandemic is both social and economic. Ethiopia, being one of the developing countries, is currently at the verge of the epidemic [3, 6, and 7].

Pregnancy is an exciting and sometimes stressful experience. Being pregnant during a disease outbreak may add extra anxiety and concern for pregnant women and for those who provide care for them. Currently, the virus is thought to be spread from an infected person to others by respiratory droplets when a person coughs or sneezes and is in close contact with another person. According to the CDC, it is not clearly known if pregnant people are more susceptible to COVID-19 than the general public. Due to changes that occur during pregnancy, pregnant people may be more susceptible to viral respiratory infections [8, 9, and 10].

There is no current evidence of adverse effects on pregnant women from COVID-19. The physical and immune system changes that occur during and after pregnancy should however be taken into account. It is critical that all women have access to safe birth, the continuum of antenatal and postnatal care, including screening tests according to national guidelines and standards, especially in epicenters of the pandemic, where access to services for pregnant women, women in labour and delivery, and lactating women is negatively impacted [11, 12].

Dealing with COVID-19 is likely to create imbalances in health care provision, disruption of routine essential services and to require redeployment of scarce health personnel across health services. Acute and emergency maternal and reproductive health services may be hit hardest, with limited facilities for isolation areas to assess and care for women in labour and the newborn [13, 14].

Pregnant women face special challenges because of their responsibilities in the workforce, as caregivers of children and other family members, and their requirements for regular contact with maternity services and clinical settings where risk of exposure to infection is higher. Due to this reason, this study tried to assess the impacts of COVID-19 infection on antenatal care services.

## 2. Objective

The general objective of this study was to assess the impact of COVID-19 infection on maternal and reproductive health care services among mothers getting service in governmental health institutions of Dessie town, 2020 G.C.

## 3. Methods and Materials

### 3.1. Study Design and Period

Institution based cross sectional study design using mixed (quantitative supplemented with qualitative) method was employed to identify the impact of COVID-19 infection on maternal and reproductive health care services among women who get service in governmental health institutions of Dessie town. The study was conducted from July 1-15 / 2020.

### 3.2. Population

The source populations were all women who get maternal and reproductive health care services in governmental health institutions of Dessie town. All women who get maternal and reproductive health care services in governmental health institutions of Dessie town during the data collection period were taken as study population.

### 3.3. Sample Size determination

The sample size in this study was determined by using single population proportion formula. Since there was no any previous study conducted on the topic, the prevalence of COVID-19 infection was taken as 50%.

The required sample based on the usual formula was as follows:

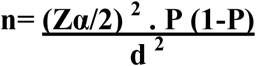

**Where** n = desired sample

P = proportion of COVID-19 infection on pregnant women to be 0.5

Zα /2 = Z value to 95% significance level=1.96

d = Margin of error (0.05) tolerated

Then, 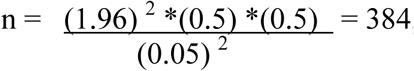, from undefined population.

n = 384, adding 10 % non-response rate, the final sample taken was **∼422**.

### 3.4. Sampling technique

Study subjects were selected from each health institution by using systematic random sampling until the minimum calculated sample size is obtained. The number of subjects selected from different hospitals and/or health centers were determined proportionally. For qualitative part a total of 20 homogeneous study subjects were selected purposely from each health institution (approximately 3 interviewees from each health institution).

### 3.5. Data collection methods and instruments

For the quantitative part, pretested and semi-structured questionnaire was used to collect the data. The data was collected by using face to face interview technique from the mothers. Questionnaires for each item were adopted from previously done similar studies and modified according to the objective of this study.

For the qualitative part, face to face in depth interview was used. During interview, the responses were recorded and the interviewer has taken notes.

### 3.6. Data quality control

From the very beginning, data collectors and supervisors have given a full course of training regarding the basic principles of data collection procedures for both quantitative and qualitative techniques. The data collection tool was pretested with 5% of the study subjects. The participants for the pretest were taken from Kombolcha 03 health center to prevent information contamination. The principal investigator and supervisors have made a day to day on site supervision during the whole period of data collection. At the end of each day, the questionnaires were reviewed and checked for completeness, accuracy and consistency by the supervisors and investigators and corrective discussion was undertaken with all the research team members.

### 3.7. Data processing, interpretation and narration

The questionnaire was checked for completeness and consistency and entered and edited in the computer for statistical analysis. Data was entered in to Epi data 4.4.1 database. Furthermore, the data editing and clearance was done on the same software. Finally, the data was taken to SPSS version 23.0 for the final analysis. The findings of the study was summarized and presented using tables, descriptive measures and statistical diagrams. The findings from the qualitative part were presented by narration.

### 3.8. Ethical consideration

The study was approved by the Ethical Review Board of Wollo University, research and community service vice president office and a cooperation letter was obtained from South Wollo health office and each respective hospital administrative office. Verbal consent was obtained from each study participants. The right of the respondents to refuse to answer for any or all questions was respected.

Names of the clients was not recorded in the questionnaires and strict confidentiality was assured through anonymous recording and coding of questionnaire and by placing them in safe place after they had been collected; and was used for the purpose of the study only.

## 4. RESULTS

### 4.1. Socio-demographic Characteristics

A total of 422 participants were responded to all the questions making the response rate 100%. Thirty seven percent of respondents were in the age group of 25-29 years. Most of the respondents were married which accounts, 346(82%). Regarding occupational status of participants, nearly half, 203 (48.1%) of them were housewives. Majority of the respondents 403(95.5%) were Amhara in ethnicity. Concerning religion of respondents, more than half of them were Muslims followed by Orthodox Christians which accounts, 262(62.1%) and 151(35.8%) respectively. Furthermore, 120(28.4%) of participants attended their secondary education and 186(44.1%) of them had higher family monthly income. (Table 1)

**Table 1:**
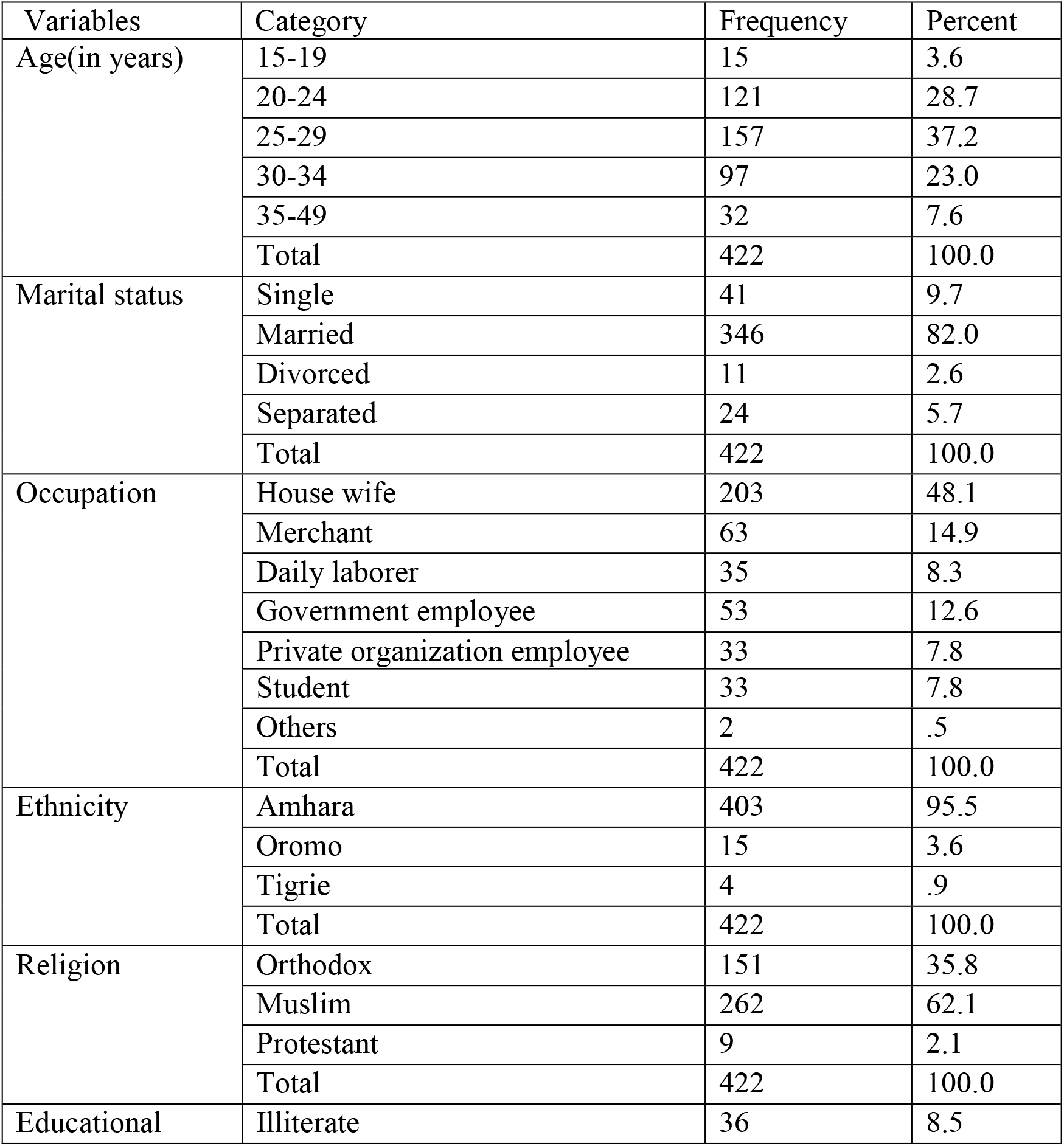
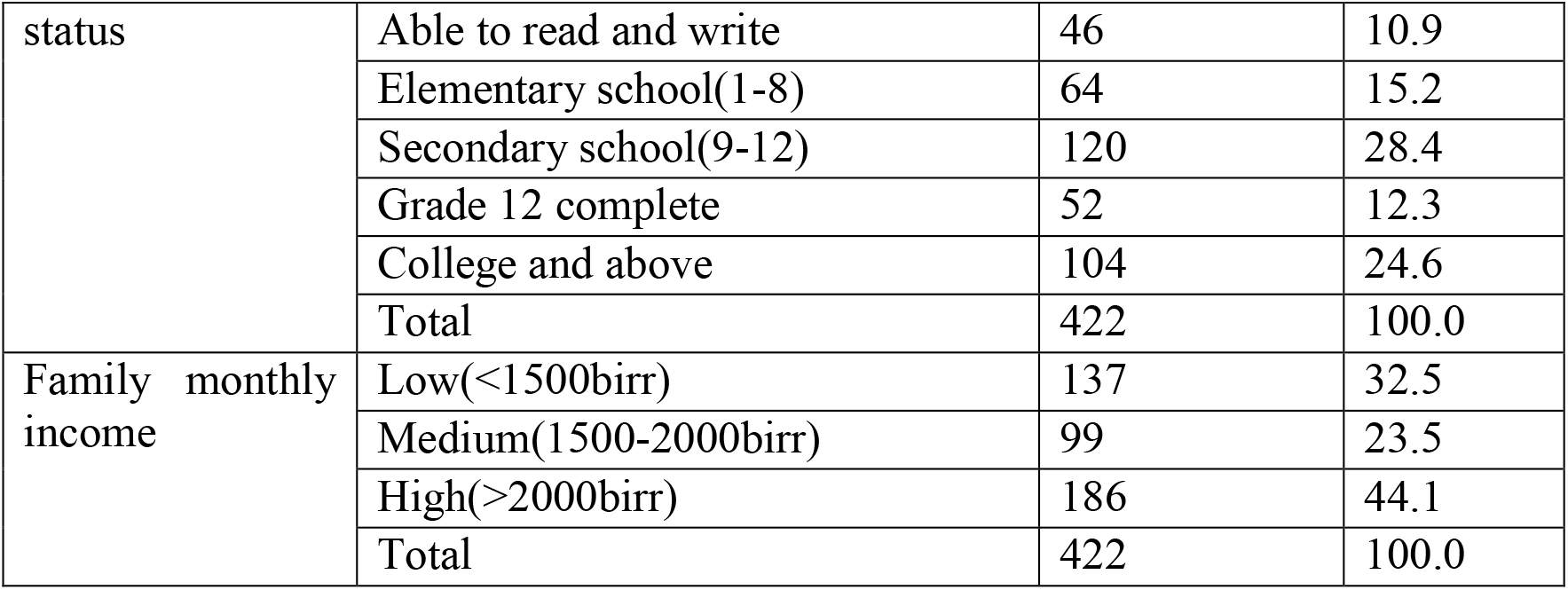
Socio-demographic and economic characteristics of study participants in Dessie governmental health institutions, 2020 (n=422).

### 4.2. Impact of covid-19 on antenatal care services

A total of 200 antenatal care attendees were asked their reasons to come to health institutions. Three-fourth of participants, 150(75%) responded that they came for regular antenatal care checkup. From those who came for regular ANC checkup, 75(50%) and 35(23.3%) of women had made their first and fourth visits respectively. From a total of 200 antenatal care attendees, 188(94%) of them reported that they had got appropriate care while 12(6%) of respondents reported that they didn’t get appropriate care. Regarding the reasons for not getting appropriate ANC care, almost all of them mentioned fear of health care providers to provide care due to Covid-19.

More than one-third (36.5%) of antenatal care attendees reported that Covid-19 affected ANC service by shifting human resource to corona prevention while nearly one-third (33.5%) of them said that COVID-19 causes cancellation of regular antenatal care visits.

The majority, 181(90.5%) of ANC attendees did not experience any changes concerning their plan of birth. Even though more than half (67%) of ANC attendees reported that nothing had changed towards prenatal care they received from health care providers, significant number of respondents(19.5%) said that the prenatal care they received from health care providers was somewhat worsened. (Refer table 2)

**Table 2:**
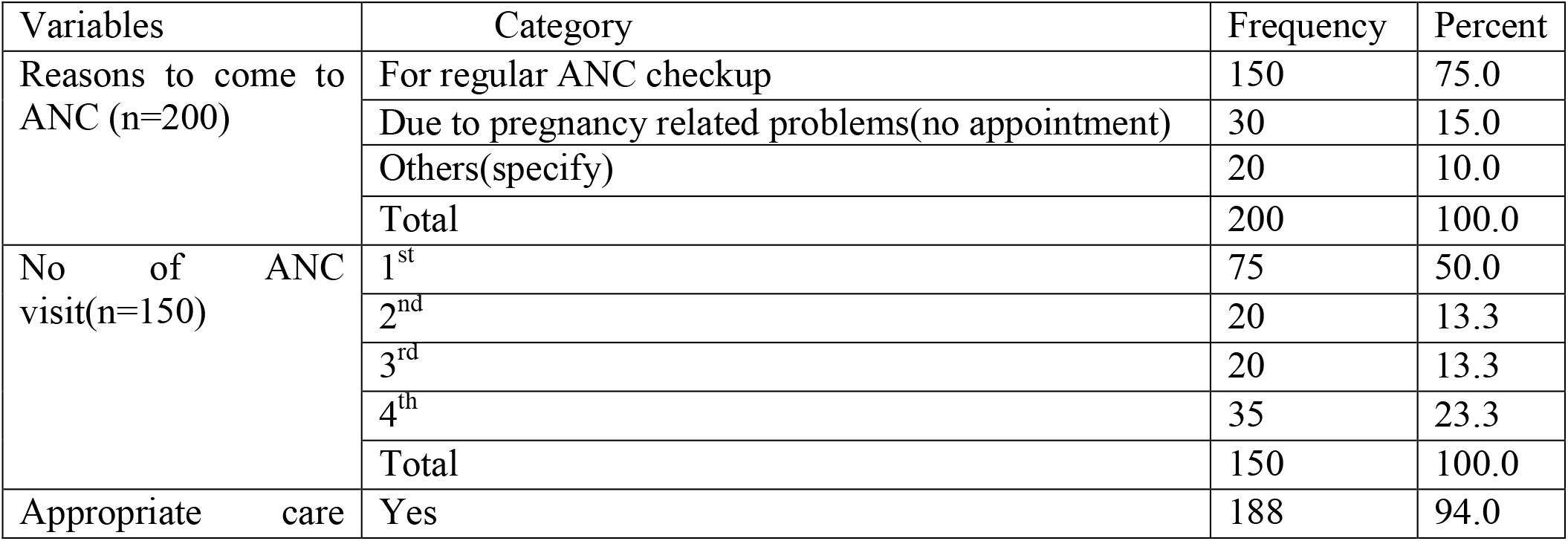
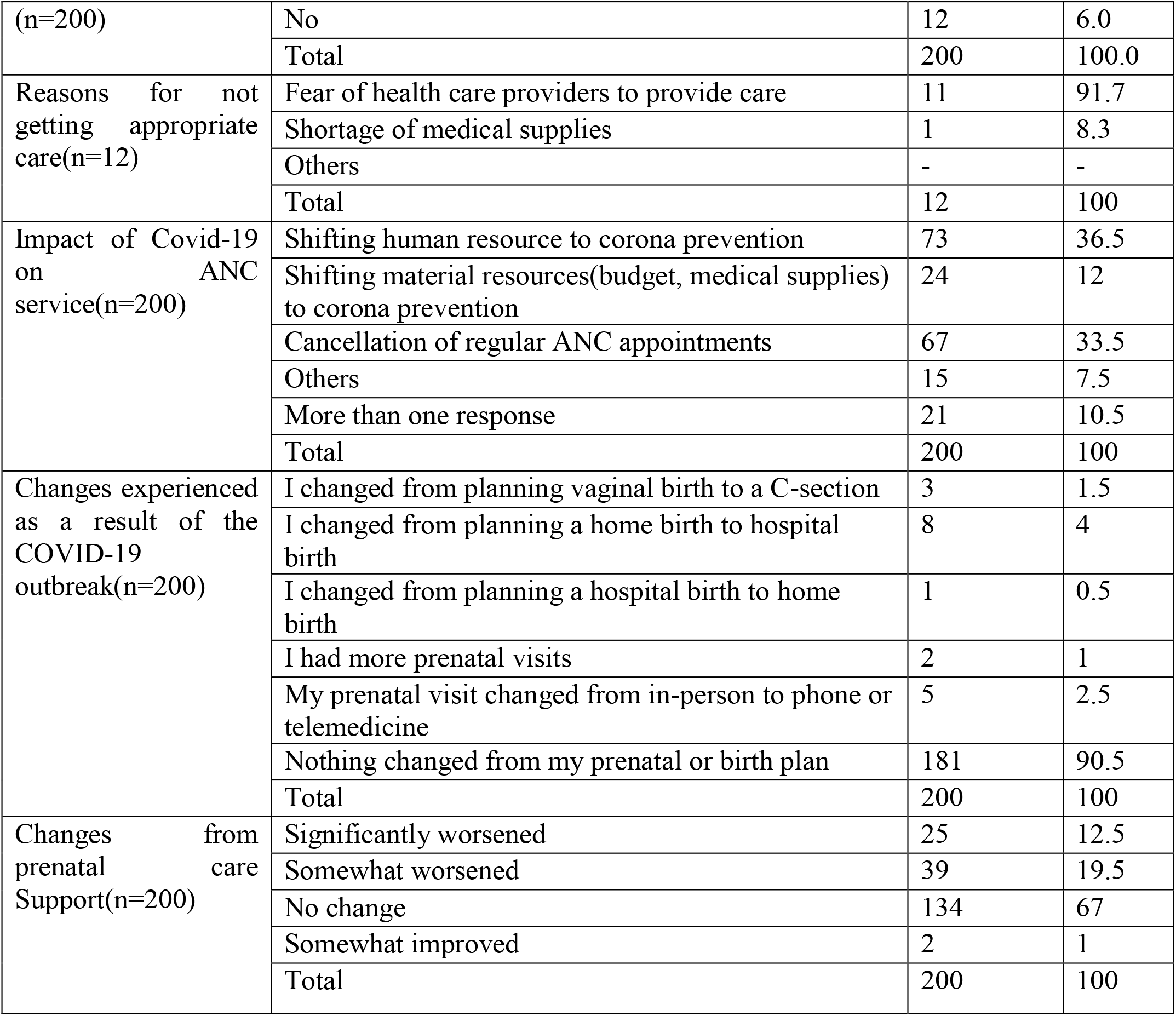
Impact of Covid-19 on antenatal care service in Dessie governmental health institutions, 2020 (n=200).

### 4.3. Impact of covid-19 on labor and delivery care services

Eighty two percent of respondents reported that they had got appropriate care while 27(18%) said that they did not get appropriate care after corona outbreak. Concerning the reasons why they did not get appropriate care, more than three-fourth (77.8%) of participants mentioned fear of health care providers. The majority, 81.3% of respondents believed that COVID-19 does not strengthen home delivery. More than one-third (39.3%) of participants reported that their support people were not allowed them to attend delivery at health institutions. Nearly one-third (29.3%) of study participants believed that the support they got from health care providers was somewhat worsened.

### 4.4. Impact of covid-19 on postnatal care services

A total of one hundred fifty (150) respondents gave birth at health institution during the data collection period. Nearly half (50.7%) of postnatal care attendees stayed at health facility greater than or equal to 24 hours while significant number(49.3%) of participants were sent to home less than 24 hours. Shortage of health care providers was the most frequently (40.5%) reported reason to send to home at less than 24 hours followed by clients desire to go to home (36.5%). More than half (53.3%) of respondents reported that they were visited by health care providers every 15 minute. Among 70(46.7%) of participants who said that they were monitored by health care providers less frequently (every 30 minute or every 1 hour), 71.4% reported that health care providers were busy. Half (50%) of postnatal care attendees said that COVID-19 had no impact on their regular child care while more than one-fourth (26.7%) of respondents had experienced difficulty of arranging their regular child care. Three-fourth (75.3%) of postnatal care attendees had changed their plan from formula feed to fully breast feeding. More than half (62%) of study participants said that the care they got from health care providers was not changed while one-third (32.7%) of them reported that the support they got from health care providers was somewhat worsened. Nearly half (49.3%) and 23.3% of respondents were mildly and extremely stressed due to the change in their newborn plans respectively. Forty two (42%) and 12.7% of respondents were stressed due to their health concerns and social distancing respectively while 36.7% cope their stress by watching television. (Refer table 4)

**Table 3:** Impact of Covid-19 on labor and delivery health care services in Dessie governmental health institutions, 2020 (n=150).

| Variables | Category | Frequency | Percent |
| --- | --- | --- | --- |
| Appropriate care (n=150) | Yes | 123 | 82 |
|  | No | 27 | 18 |
|  | Total | 150 | 100.0 |
| Reasons for not getting appropriate care(n=27) | Fear of health care providers to provide care | 21 | 77.8 |
|  | Shortage of health care providers | 3 | 11.1 |
|  | Shortage of medical supplies | 3 | 11.1 |
|  | Total | 27 | 100 |
| COVID-19 strengthens home delivery | Strongly agree | 4 | 2.7 |
|  | Agree | 6 | 4 |
|  | Disagree | 122 | 81.3 |
|  | Strongly disagree | 18 | 12 |
|  | Total | 150 | 100 |
| Changes experienced as a result of the COVID-19 outbreak(n=150) | My support people(spouse/partner, family) were not permitted to attend delivery at health institution | 59 | 39.3 |
|  | I was separated from my baby immediately after delivery | 47 | 31.3 |
|  | Other | 42 | 28 |
|  | More than one response | 2 | 1.3 |
|  | Total | 150 | 100 |
| Support changes from health care Support(n=150) | Significantly worsened | 9 | 6 |
|  | Somewhat worsened | 44 | 29.3 |
|  | No change | 88 | 58.7 |
|  | Somewhat improved | 6 | 4 |
|  | Significantly improved | 3 | 2 |
|  | Total | 150 | 100 |

**Table 4:** Impact of Covid-19 on postnatal care service in Dessie governmental health institutions, 2020 (n=150).

| Variables | Category | Frequency | Percent |
| --- | --- | --- | --- |
| Duration of stay at health facility (n=150) | <24hours | 74 | 49.3 |
|  | >=24hours | 76 | 50.7 |
|  | Total | 150 | 100.0 |
| Reasons for staying less than 24 hours (n=74) | Shortage of beds | 17 | 22.9 |
|  | Shortage of health care providers | 30 | 40.5 |
|  | I want to go home | 27 | 36.5 |
|  | Total | 74 | 100.0 |
| Frequency of visit by health care provider immediately after birth (n=150) | Every 15 minutes | 80 | 53.3 |
|  | Every 30 Minute | 43 | 28.7 |
|  | Every one hour | 27 | 18 |
|  | Total | 150 | 100 |
| Reasons for less frequent visit by health care providers(n=70) | Shortage of health care providers who can provide postnatal care | 14 | 20 |
|  | Health care providers were busy | 50 | 71.4 |
|  | Health care providers negligence | 6 | 8.6 |
|  | Total | 70 | 100 |
| Impact of Covid-19 on regular child care (n=150) | I had difficulty of arranging regular childcare | 40 | 26.7 |
|  | I had to pay more for childcare | 13 | 8.6 |
|  | My spouse/partner or I had to change our work schedule to care for our children ourselves | 18 | 12 |
|  | My regular childcare has not been affected by COVID-19 outbreak | 75 | 50 |
|  | I don't have a child in childcare | 4 | 2.7 |
|  | Total | 150 | 100 |
| Changes experienced due to COVID-19 (n=150) | I changed from planning to breastfeed to feeding only formula | 16 | 10.7 |
|  | I changed from planning to feed only formula to breastfeeding | 113 | 75.3 |
|  | Nothing changed in my newborn plans | 21 | 14 |
|  | Total | 150 | 100 |
| Changes from postnatal Support(n=200) | Significantly worsened | 7 | 4.7 |
|  | Somewhat worsened | 49 | 32.7 |
|  | No change | 93 | 62 |
|  | Somewhat improved | 1 | 0.6 |
|  | Total | 150 | 100 |
| Stress due to changes in newborn care plan due to the COVID19 outbreak | Not at all | 22 | 14.7 |
|  | Mildly | 74 | 49.3 |
|  | Moderately | 19 | 12.7 |
|  | Extremely | 35 | 23.3 |
|  | Total | 150 | 100 |
| Source of stress from COVID-19 | Health concerns | 63 | 42 |
|  | Financial concerns | 11 | 7.3 |
|  | Impact on work | 9 | 6 |
|  | Impact on your child | 11 | 7.3 |
|  | Impact on your community | 5 | 3.3 |
|  | Impact on family members | 12 | 8 |
|  | Access to food | 3 | 2 |
|  | Access to baby supplies (e.g., formula, diapers, wipes) | 7 | 4.7 |
|  | Access to personal care products or household supplies | 2 | 1.3 |
|  | Access to medical care, including mental health care | 7 | 4.7 |
|  | Social distancing or being quarantined | 19 | 12.7 |
|  | I am not stressed about the COVID-19 outbreak | 1 | 0.7 |
|  | Total | 150 | 100 |
| Things done to cope with stress due to COVID-19 | Talking with friends and family (e.g., by phone, text, or video) | 19 | 12.7 |
|  | Engaging in more family activities (e.g., games, sports) | 31 | 20.7 |
|  | Increased television watching or other activities (e.g., video games, social media) | 55 | 36.7 |
|  | Eating more often, including snacking | 5 | 3.3 |
|  | Increasing time reading books, or doing activities like puzzles and crosswords | 18 | 12 |
|  | Talking to my healthcare providers more frequently, including mental healthcare provider (e.g., therapist, psychologist, | 7 | 4.7 |
|  | Volunteer work | 4 | 2.7 |
|  | I have not done any of these things to cope with the COVID-19 outbreak | 10 | 6.7 |
|  | Total | 150 | 100 |

### 4.5. Impact of covid-19 on reproductive health care services

A total of 72 clients were asked regarding their reproductive health care service utilization after corona outbreak. More than half (65.3%) of respondents had ever used contraceptives after corona outbreak. More than one-third (34.7%) of the study participants did not ever used contraceptives due to unavailable service (32%), travel restrictions (28%) and no plan to use family planning(40%). Majority (90.3%) of respondents did not experience abortion after corona outbreak while only 9.3% of them had experienced it. From those who had experienced abortion, more than half (57.1%) of them did not go to health institutions since they think that the service may not be given as a result of corona. Most of the respondents (84.7%) did not experience unwanted pregnancy after COVID outbreak. Among those who had experienced unwanted pregnancy, more than half (63.6%) of them were due to their thought that they can’t get family planning service due to corona. (Refer table 5)

**Table 5:** Impact of Covid-19 on postnatal care service in Dessie governmental health institutions, 2020 (n=72).

| Variables | Category | Frequency | Percent |
| --- | --- | --- | --- |
| Ever use of contraceptives after corona outbreak (n=72) | Yes | 47 | 65.3 |
|  | No | 25 | 34.7 |
|  | Total | 72 | 100.0 |
| Reasons for not using contraceptives(n=25) | The service is not available due to corona outbreak | 8 | 32 |
|  | Due to travel restriction | 7 | 28 |
|  | I have no plan to use family planning | 10 | 40 |
|  | Total | 25 | 100 |
| Abortion experience after corona outbreak(n=72) | Yes | 7 | 9.7 |
|  | No | 65 | 90.3 |
|  | Total | 72 | 100 |
| Go to health institutions to get abortion service (n=7) | Yes | 3 | 42.9 |
|  | No | 4 | 57.1 |
|  | Total | 7 | 100 |
| Reasons for not going health institution(n=4) | I think I can't get the service due to corona | 4 | 100 |
|  | I have used herbal drugs to cause abortion | 0 | - |
|  | Total | 4 | 100 |
| The way health care providers treated you | The HCP told me that the service is not available at the moment | 2 | 50 |
|  | The HCP treated me as usual | 2 | 50 |
|  | Total | 4 | 100 |
| Experience of unwanted pregnancy after corona outbreak(n=72) | Yes | 11 | 15.3 |
|  | No | 61 | 84.7 |
|  | Total | 72 | 100 |
| Reasons for occurrence of unwanted pregnancy(n=11) | I can't get family planning service due to corona | 7 | 63.6 |
|  | Unprotected sex due to shortage of condom | 3 | 27.3 |
|  | Failure of contraceptives | 1 | 9.1 |

### 4.6. Pregnancy, delivery, postnatal and reproductive health history related data (Document review)

The document review indicated that the number of clients who came to health institutions to get different maternal and reproductive health care services after outbreak of COVID-19 in Ethiopia (March to June) was decreased. The numbers of 2nd and 3rd ANC visits were decreased significantly, 83.8% and 88.2% respectively. Regarding the rout of delivery, the number of women who gave birth by c/s was increased by 8%. The number of maternal and neonatal death was also increased significantly, 16.7% and 28.6% respectively. Concerning contraceptive use, utilization of all forms contraceptives was decreased. (Refer table 6)

**Table 6:** Pregnancy, delivery, postnatal and reproductive health history related data (Document review)

| S.No | Variable | Category | November to February | March to June | Total | Variation (in %) |
| --- | --- | --- | --- | --- | --- | --- |
| 1. | Number of ANC visits | 1<sup>st</sup> | 480(96.6%) | 317(63.9%) | 497 | 32.7 |
|  |  | 2<sup>nd</sup> | 137(91.9%) | 12(8.1%) | 149 | 83.8 |
|  |  | 3<sup>rd</sup> | 64(94.1%) | 4(5.9%) | 68 | 88.2 |
|  |  | 4<sup>th</sup> | 235(67.5%) | 113(32.5%) | 348 | 35 |
| 2. | Rout of delivery | SVD | 1515(50.3%) | 1497(49.7%) | 3012 | 0.6 |
|  |  | Instrumental | 98(56%) | 77(44%) | 175 | 12 |
|  |  | Cesarean section | 444(46%) | 521(54%) | 965 | 8(increment) |
| 3. | History of death | Still birth | 51(64.6%) | 28(35.4%) | 79 | 29.1 |
|  |  | Maternal death | 5(41.7%) | 7(58.3%) | 12 | 16.7(increment) |
|  |  | Neonatal death | 80(35.7%) | 144(64.3%) | 224 | 28.6(increment) |
| 4. | Contraceptive use | Pills | 101(72.7%) | 38(27.3%) | 139 | 45.3 |
|  |  | Condom | 5(100) | 0 | 5 | 100% |
|  |  | Injectable | 324(55.9%) | 256(44.1%) | 580 | 11.7 |
|  |  | IUCD | 169(57.9%) | 123(42.1%) | 292 | 15.7 |
|  |  | Implants | 504(65.5%) | 265(34.5%) | 769 | 31.1 |
|  |  | Permanent | 0 | 0 | 0 | - |

### 4.7. Qualitative result

For the qualitative part, 20 participants were interviewed by face to face in depth interview. During interview, the responses were recorded and the interviewers have taken notes. The responses are summarized in 5 sections.

#### 4.7.1. Impact of COVID-19 infection on pregnancy

Most study participants were stressed due to risk of corona transmission from health care providers while they provide care for them. They also described that they were stressed because they think that if they became infected with corona it may affect their fetus. A 27-year-old woman said *‘‘I came to the hospital for my first antenatal care and asked the doctor the impact of corona on my fetus’’*.

A 34-year-old woman said *‘‘I came here for my 4*^*th*^ *visit due to the influence of my family because I fear to contract corona virus from other clients as well as from health care providers*.*’’*

#### 4.7.2. Impact of COVID-19 infection on Antenatal care

All of the study participants described that corona virus has significant negative impact on the quality of antenatal care. It causes cancellation of 2^nd^ and 3^rd^ antenatal visits. A 30-year-old woman reported that *‘‘I came to the health center for my 4*^*th*^ *antenatal checkup, but the health care provider didn’t touch my abdomen, not even check my fetal movement*.*’’*

*Another 26-year-old woman said that ‘‘I came here for my 1*^*st*^ *antenatal visit, the health care provider ordered me to have urine test and told me that I am pregnant. He didn’t tell me even what I should do. When I asked him when to come for my next visit, he said that no need to come for further checkup unless labor comes*.*’’*

#### 4.7.3. Impact of COVID-19 infection on delivery care

Most of interviewees expressed that health care providers didn’t frequently visit them due to shortage of health care providers. *A 33-year-old woman said that ‘‘I was in labor ward. My labor pain became worse and I was shouting and calling the doctor to see me, but no one treated me for at least 2 hours after admission*.*’’*

Some of the interviewees described that the care they got from health care providers didn’t change due to corona; rather the care was improved a little bit. *A 35-year-old woman described her experience as ‘‘I was admitted in labor ward and the health care providers provided me special care better than usual; all of them wore masks &gloves and has washed their hands frequently*.*’’*

#### 4.7.4. Impact of COVID-19 infection on postnatal care

Some interviewees reported that the postnatal care provided was significantly decreased. After delivery health care providers didn’t visit frequently and sent clients to home at less than 24 hours of hospital stay. *A 25-year-old woman said that ‘‘I had faced difficulty of breast feeding after birth and no one helped me to breast feed my neonate. I was also sent to home after 6 hours of hospital stay*.*’’*

#### 4.7.5. Impact of COVID-19 infection on reproductive health care

One of the health care services negatively affected by corona was reproductive health care as the concern of the health sector health force is shifted towards corona prevention. *A 28-year-old divorced women reported that ‘‘my sister has gone to health institution to get family planning service after corona outbreak in Ethiopia. The health care provider told her that it is not good to come to health institution at this time unless severely ill. You can buy pills at pharmacy or communicate your doctor with phone calls*.*’’*

*A 27-year-old lady also said that ‘‘I fear to go to health institutions to get reproductive health care services because I think the health institutions is full of corona. Rather we prefer to purchase drugs at pharmacies. I know a woman who has experienced abortion following unwanted pregnancy after corona outbreak, but she didn’t go to health institution due to fear of contracting corona*.*’’*

## 5. Discussion

This study tried to assess the impact of COVID-19 on maternal and reproductive health care services in governmental health institutions of Dessie town. The result of this study was discussed with different findings all over the world.

Even though the majority of antenatal care attendees said that they got appropriate antenatal care, some respondents reported inappropriate care due to fear and shortage of health care providers which may strengthen home delivery. The finding of this study is in line with the finding in America which reported that an increasing number of health workers are being exposed, while midwifery services are overwhelmed by calls from concerned mothers now exploring home birth options. The problem of shortage of health care providers might be more profound in developing countries like Ethiopia [6, 7].

According to this study, three-fourth (74.5%) of ANC attendees agreed that covid-19 decreases ANC visits. The finding of this study revealed that the most frequent reported impact of Covid-19 on ANC service were shifting of human resources to corona prevention (36.5%) followed by cancellation of ANC visits (33.5%). Similarly, COVID-19 Technical Brief Package for Maternity Services reported that Deploying maternity care workers away from providing maternity care to work in public health or general medical areas during this pandemic is likely to increase poor maternal and newborn outcomes. Ten percent (10%) decline in service coverage of essential pregnancy-related and newborn care (1,745,000 additional women experiencing major obstetric complications without care) were reported from 132 low- and middle-income countries. This similar finding could be justified that the impact of COVID-19 is global [22, 23].

In this study, eighteen percent (18%) of delivery care attendees reported that they didn’t get appropriate care, of this 35.3% of them said that the support they got from health care providers was worsened. The result is similar with the study conducted by Ashish KC, Rejina G and Mary V which indicated that delivery care after the pandemic was decreased (health workers greeting the mother decreased by 2.2% (–3.1 to –1.3), the use of gloves and gown for childbirth decreased by 2.4% (–3.1 to –1.9), companionship during labour decreased by 6.0% (–6.9 to –5.1), and intrapartum fetal heart rate monitoring decreased by 13.4% (–15.4 to –11.3). This might be due to fear of health care providers to frequently make contacts with laboring mother and shortage of medical supplies such as gloves [24].

This study revealed that nearly half (49.3%) of postnatal care attendees reported that they stayed at health facility less than 24 hours after delivery and the most frequently mentioned reason to stay at less than 24 hours was clients desire to go home. This finding is similar with a study conducted in Europe by Kirstie C and Cristina F which reported that women are sent home more quickly after a miscarriage or stillbirth, and accounts of bereavement rooms being re-allocated to the care of women who have COVID-19 [21]. The reason might be clients’ attitude that staying more at health facility could increase risk of acquiring COVID-19,

In this study, nearly half (46.7% of respondents said that they were visited less frequently after delivery and the reason was that health care providers were busy which accounts 71.4%). Similar study conducted by Kirstie C and Cristina F in Europe showed that maternity providers have often limited follow-ups, and as in other EU countries, some UK hospitals have stopped allowing birth partners to be present[21]. The reason could be justified that maternity care providers are shifted to COVID-19 prevention as well as shortage of necessary medical supplies to frequently visit clients.

In our study, significant proportion of postnatal care attendees (37.4%) reported that the support they received from health care providers was worsened. Similarly the study in Nepal reported decreased postnatal care (placing the baby skin-to-skin with the mother decreased from 30% to 20%, and breastfeeding within 1 hour of birth decreased by 3.5% (–4·6 to –2·6). The possible explanation could be health care providers may worry that skin to skin contact and breast feeding would transmit the virus to newborns. The study in Nepal also showed that immediate newborn care practice of cord clamping 1 min after birth increased slightly after the outbreak. This could be due to health care providers’ assumption that immediate cord cut may reduce risk of transmission to the newborn [24].

In this study, among women who were in need of contraceptives, nearly one-third (32%) of them didn’t use contraceptives due to unavailability of the service. The result is in line with the study in America which reported that women were unable to access family planning and some of these women stopped going to facilities due to fear of infection and increased physical and financial barriers [9, 10]. Another study done by Taylor Riley, Guttmacher, Elizabeth Sully et al also indicated that COVID-19 pandemic is already having adverse effects on the supply chain for contraceptive commodities by disrupting the manufacture of key pharmaceutical components of contraceptive methods or the manufacture of the methods themselves (e.g., condoms), and by delaying transportation of contraceptive commodities. A report from 132 low- and middle-income countries indicated that Potential annual impacts of a 10% proportional decline in use of sexual and reproductive health care services resulting from COVID-19–related disruptions [11, 12, and 23].

In this study, more than half (57.1%) of respondents who has experienced abortion didn’t go to health institutions since all (100%) of them think that the service may not be available during this corona period. This study also showed that, among those who have experienced unwanted pregnancy after corona outbreak, 63.6% of them mentioned unavailability of family planning services as a reason for the occurrence of unwanted pregnancy. This is similar with the study done by Taylor Riley, Guttmacher, Elizabeth Sully et al which reported that and providers are being forced to suspend some sexual and reproductive health services that are not classified as essential, such as abortion care, thus denying people this time-sensitive and potentially life-saving service. Ten percent (10%) shifts in abortions from safe to unsafe (3,325,000 additional unsafe abortions) were reported from 132 low- and middle-income countries [14, 15, and 23].

In this study, a substantial decrease in institutional delivery was seen (in spontaneous vaginal delivery and instrumental delivery, 0.6% and 12% respectively) while the number of women who gave birth by c/s was increased by 8% (from 46% to 54%). Similar study conducted by Ashish KC, Rejina G and Mary V revealed that a substantial decrease (7.4%) in institutional delivery was seen between January and May, 2020. The same study also revealed that the proportion of women who had caesarean section increased from 24.5% (n=3234) before lockdown to 26.2% (n=1879) during lockdown (p=0·0075). The reason might be due to that women came to heath institutions for delivery only if they faced complications otherwise they would give birth at home during this pandemic [24].

The neonatal death in this study was increased from 35.7% to 64.3%. This finding is in line with the study done in Nepal which reported that the institutional neonatal mortality rate increased from 13 deaths per 1000 live births before lockdown to 40 deaths per 1000 live births during the lockdown. This might be due to decreased quality and frequency of antenatal care visits as well as maternal stress due to COVID-19. The current study showed a decreased still birth from 64.6% to 35.4%. In contrast to this, the study in Nepal revealed an increased in still births from 14 to 21/1000 total births. This might occur due to difference in sample population, study period as well as study design [24].

In this study, the proportion of maternal complications was increased from 41.7% to 58.3% which was higher than the finding in Nepal in which the proportion of women who had a complication was increased from 6.7% (n=884) before lockdown to 8.7% (n=587) during lockdown (p=0·0126). The difference might be justified that the quality of health care was better in Nepal (the supply of medical equipment’s that help to prevent COVID-19 in Ethiopia was low [24].

## 6. Conclusion and recommendation

### 6.1. Conclusion

This study concluded that COVID-19 significantly affects the quality and utilization of maternal and reproductive health care services. According to EDHS 2016 report, the utilization of maternal and reproductive health care services in Ethiopia was low (institutional delivery-26%, antenatal care coverage-34% and contraceptive prevalence rate-36%) and Covid-19 could further decrease this coverage.

According to this study, Six percent (6%) of antenatal care attendees, 18% of delivery care attendees and nearly half (46.7%) of postnatal care attendees reported inappropriate service delivery due to fear of health care providers, shortage medical supplies and staff work load. The study also showed that utilization of these services was decreased due to fear of clients to go to health institutions.

Significant number (32% of antenatal care attendees, 35.3% of delivery care attendees and 37.4% of postnatal care attendees reported that the Support they received from the health care provider was worsened due to COVID-19. Additionally, more than one –third (33.5%) of antenatal care attendants reported cancellation of regular ANC visits.

The majority (85.3%) of participants was stressed due to corona and health concern was the greatest source of stress which accounts 42%.

Most (93.3%) of the respondents reported that COVID-19 has negative impact on their life, of which nearly one-third (29.3%) of them stated it as extremely negative impact.

Significant numbers of respondents have experienced unwanted pregnancy after corona outbreak. Of these, 63.6% of them mentioned unavailability of family planning services as a reason for the occurrence of unwanted pregnancy.

The qualitative finding indicated that Covid-19 has significant negative impact on maternal and reproductive health care services (stress, cancellation of ANC visits, decreased quality and no desire to go to health institutions as clients think health institutions is risky place).

The data collected from documents indicated that the number of clients who have used different maternal and reproductive health care services was significantly decreased.

### 6.2. Recommendation

According to the findings of this study, the researchers would like to forward the following recommendations.

➢ **To ministry of health:** ministry of health should
  ✥ Continue maternity services to be prioritized as an essential core health service, and other sexual and reproductive health care such as family planning, emergency contraception, treatment of sexually transmitted infections, and where legal safe abortion services, to the full extent of the law, also need to remain available as core health services.
  ✥ Provide full access of all personal protective equipment (PPE), sanitation and a safe and respectful working environment for maternity care providers
➢ **To health institutions:** Health institutions should
  ✥ Avail all necessary medical supplies (gloves, masks, sanitizer etc) for both health care providers and clients to improve quality of maternal and reproductive health care services.
  ✥ Assign adequate number of health care providers at each unit/ward.
  ✥ Provide health education for clients to avoid misunderstanding towards corona as respondents think going to health institution increases risk of acquiring it.
  ✥ Improve the quality of their services to attract clients
➢ **To Wollo University:** Wollo University should
  ✥ Organize awareness creation program at community level to increase utilization of maternal and reproductive health care services.
➢ **To researchers**
  ✥ Further study using strong study design with additional variables should be conducted in the future.

**Fig 1:**
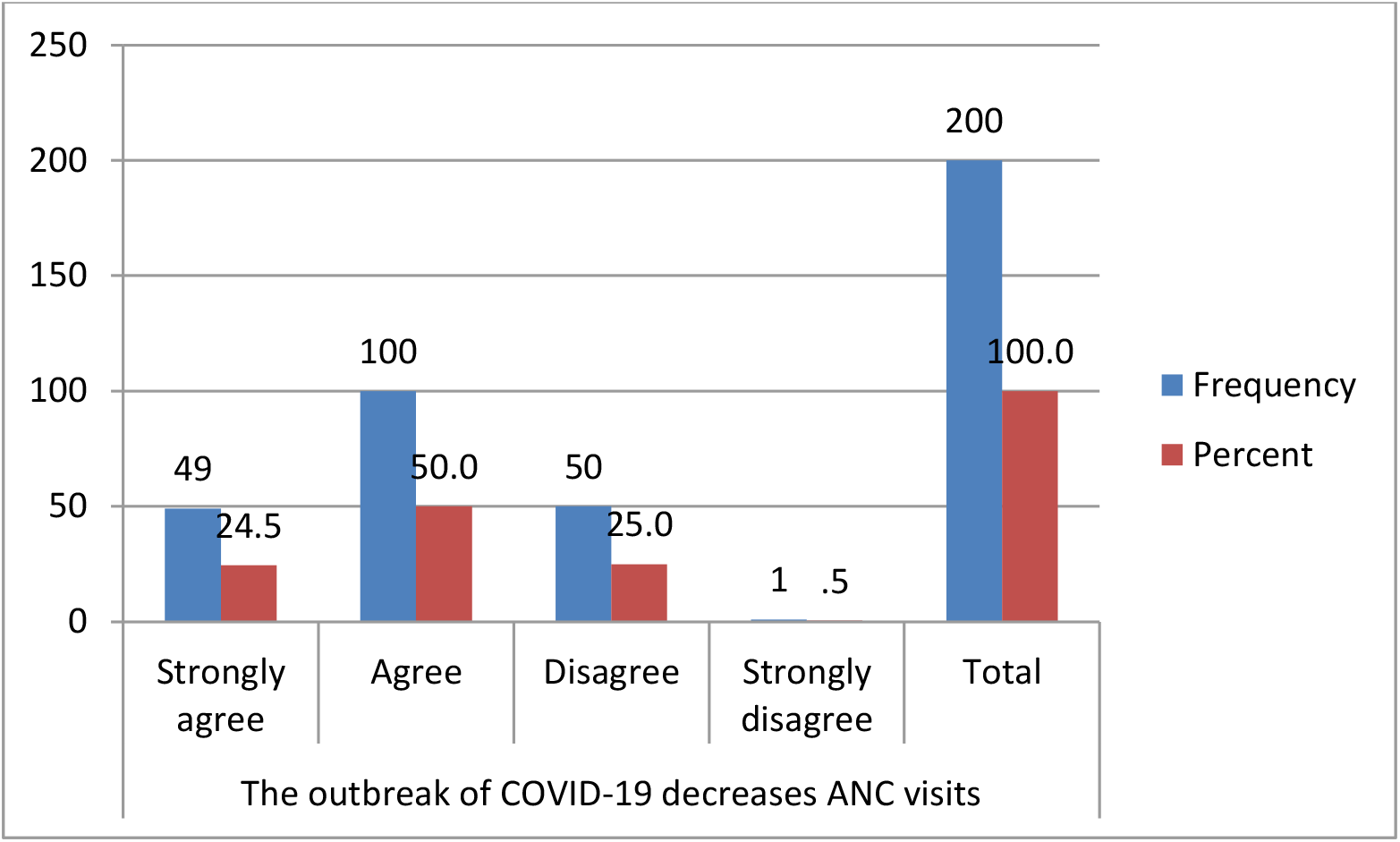
Impact of COVID-19 on ANC visit. Figure 1 indicated that Half (50%) and nearly one-fourth (24.5%) of antenatal care attendees agree and strongly agree with the statement COVID-19 decreases ANC visits respectively.

**Fig 2:**
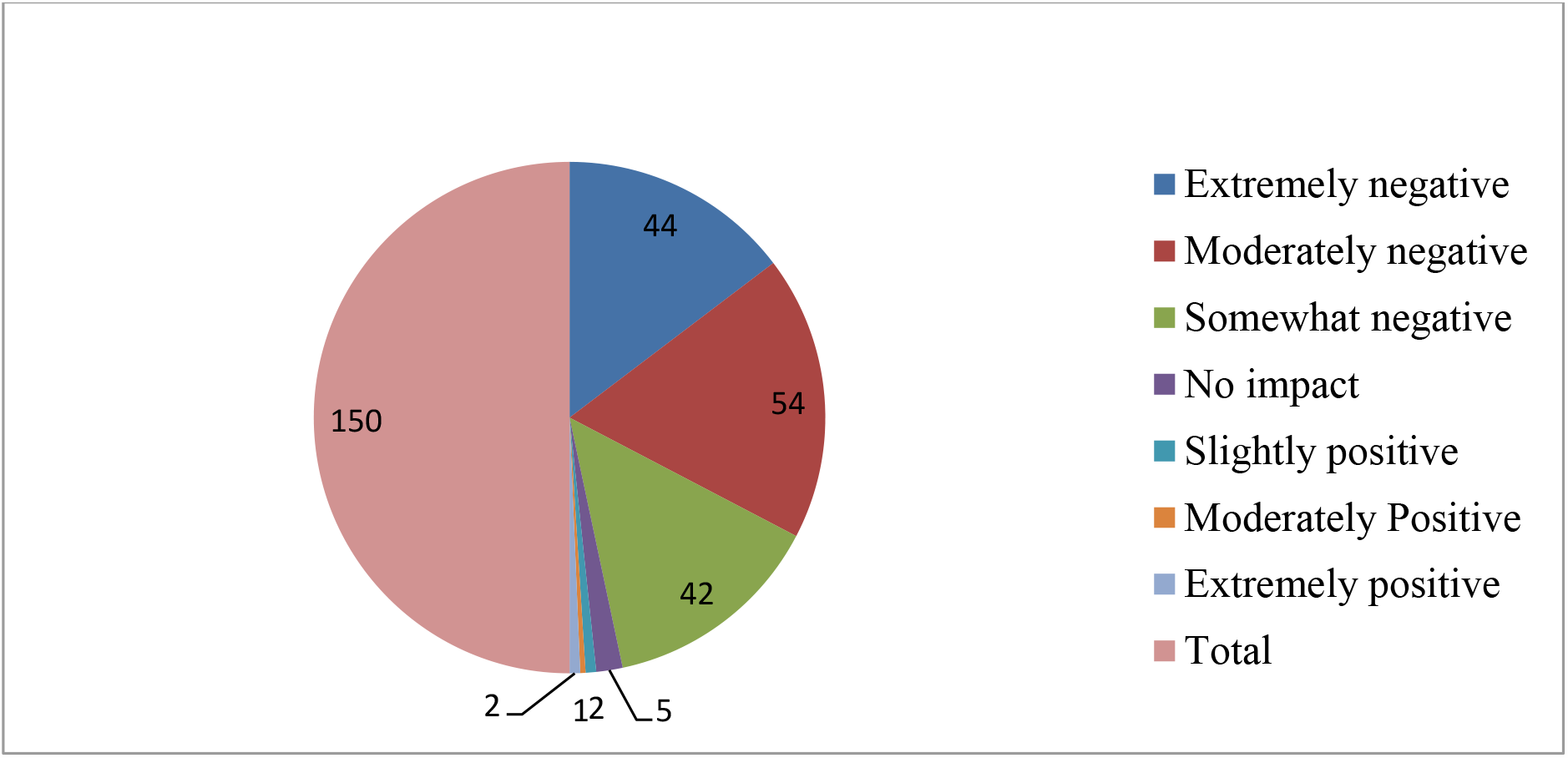
Extent of the impact of COVID-19 on participants life. Thirty six percent (36%) of respondents reported that COVID-19 has moderately negative impact on their life while nearly one-third (29.3%) of postnatal care attendees said that COVID-19 has extremely negative impact on their life. (Refer fig 2)

## Data Availability

All the data needed are available

https://www.guttmacher.org

## ABBREVIATIONS

CDC: Communicable disease control
COVID-19: Corona virus disease 2019
ICTV: International committee on taxonomy of virus
MERS: Middle East respiratory syndrome
MMR: Maternal Mortality Rate
nCOV: Novel corona virus
RNA: Ribonucleic acid
SARS: Sever acute respiratory syndrome
SARS-CoV-2: Sever acute respiratory syndrome-corona virus-2
WHO: World Health Organization

## DECLARATIONS

### ETHICAL APPROVAL AND CONSENT TO PARTICIPATE

The study was approved by the Ethical Review Board of Wollo University and a cooperation letter was obtained from each health institutions administrative office.

Written consent was obtained from each study participants. The right of the respondents to refuse to answer for any or all questions was respected.

### CONSENT FOR PUBLICATION

All authors fully agreed.

### AVAILIBILITY OF DATA AND MATERIALS

all data and materials for this manuscript are available

### COMPETING INTERESTS

The authors declared that there is no conflict of interest in this research article. Please contact author for data requests.

### FUNDING

All cost related to this research was covered by Wollo University research and community service coordinating office.

### AUTHORS’ CONTRIBUTIONS

The data collection was undertaken by all the authors. All the authors contributed in the data analysis, design and preparation of the manuscript. All authors read and approved the final manuscript and have all agreed to its submission for publication.

## ACKNOWLEDGEMENTS

First of all, we would like to express our sincere thanks to the Almighty God for His love, forgiveness and generosity.

It gives great pleasure to present our sincere thanks for the support we got from the administrative staffs of each governmental health institutions.

Our thanks also go to Wollo University, research and community service coordinating office for allowing us to do this research work.

## Notes

### Competing Interest Statement

The authors have declared no competing interest.

### Clinical Trial

This study is not a clinical trial.

### Funding Statement

No external fund was received.

### Author Declarations

The study was approved by the Ethical Review Board of Wollo University, research and community service vice president office and a cooperation letter was written to each health institutions.

